# Socioeconomic changes predict genome-wide DNA methylation in childhood

**DOI:** 10.1101/2021.06.23.21259418

**Authors:** Jiaxuan Liu, Janine Cerutti, Alexandre A. Lussier, Yiwen Zhu, Brooke J. Smith, Andrew D.A.C. Smith, Erin C. Dunn

## Abstract

Childhood socioeconomic position (SEP) is a major determinant of health and well-being across the entire life course. To effectively prevent and reduce health risks related to SEP, it is critical to better understand when and under what circumstances socioeconomic adversity shapes biological processes. DNA methylation (DNAm) is one such mechanism for how early life adversity “gets under the skin”. Using data from a large, longitudinal birth cohort, we showed that *changes* in the socioeconomic environment may influence DNAm at age 7. We also showed that middle childhood (ages 6-7) may be a potential sensitive period when socioeconomic instability, reflected in parental job loss, is especially important in shaping DNAm. Our findings highlight the importance of socioeconomic stability during childhood, providing biological evidence in support of public programs to help children and families experiencing socioeconomic instability and other forms of socioeconomic adversity during childhood.

## Introduction

Socioeconomic position (SEP) is a fundamental determinant of health and disease across the lifespan^1^. As defined by Krieger et al.^2^, SEP is an “aggregate concept” composed of diverse components of economic and social well-being across individual-, household-, and neighborhood-level domains, including both resources (e.g., weekly income) and rank-based characteristics (e.g., occupational prestige). SEP therefore can be measured across time by various indicators, like job stability, ability to afford basic household needs, and neighborhood quality, which are known to play related, yet distinct roles in health and life outcomes^3–5^.

Dozens of observational and quasi-experimental studies examining these indicators have shown that children growing-up in low SEP families are at increased risk for both short- and long-term cognitive, socioemotional, behavioral, and physical/mental health deficits compared to their high SEP counterparts^6–9^. Some of these SEP disparities are evident very early in development, starting shortly after birth^10^. For example, in one of the largest studies to date on SEP and health (covering 1.7 million individuals across seven high-income countries), low SEP was linked to a 2-year reduction in life expectancy, a larger effect than those observed for years-of-life lost due to obesity, hypertension, alcohol intake, and other risk factors^11^. In fact, living in socioeconomic deprivation reduces life expectancy by as much as a decade in both the United States^12^ and England^13^. Yet, little is understood about the biological mechanisms that explain these well-established SEP and health relationships, limiting our ability to disentangle specific pathways of pathophysiology and design targeted intervention.

In the past two decades, epigenetic studies have exploded as a means of potentially unraveling the biological pathways through which SEP “gets under the skin”. Most epigenetic studies have focused on DNA methylation (DNAm)^14^, which occurs when methyl groups are added to cytosines in the DNA sequence, typically within cytosine-guanine (CpG) dinucleotides^15^. These DNA modifications do not alter the sequence of the genome, but can influence how genes are expressed in ways that can have important short and long-term health consequences^16^.

Recent reviews summarizing the effects of SEP on epigenetic patterns suggest that SEP is linked to DNAm differences in childhood and adulthood^17–19^. In fact, over 30 studies have found a relationship between childhood SEP and DNAm. However, the literature is mixed, with no consistent patterns in SEP-associated DNAm changes emerging between studies. One possible explanation for these mixed results is that studies have conflated both the type of SEP indicator measured and the timing of SEP measurement^19^. Indeed, very few studies have investigated the developmental patterning of different SEP domains (e.g., household income and neighborhood) and their effects on DNAm, even though it is well known that the type and timing of SEP can influence the extent of its impact^20^.

In some notable exceptions, studies comparing the time-dependent effects of childhood SEP^21–23^ or other types of childhood adversities^24–26^ on DNAm have found timing differences with respect to SEP’s impact, consistent with the idea that there may be *sensitive periods* of elevated plasticity during childhood when adversity-induced biological changes are most likely to occur. However, whether changes in the socioeconomic environment, for better or for worse, influence the SEP-DNAm relationship across developmental stages remain unexplored. These answers are needed to develop targeted interventions and policies aimed at reducing the negative health consequences of low childhood SEP.

Here, we sought to assess how the socioeconomic environment in the first seven years of life – including socioeconomic mobility, instability, and the developmental timing of socioeconomic disruptions – associated with epigenetic alterations. Because socioeconomic adversity could have multiple time-varying effects on DNAm, we tested three commonly examined hypotheses from the life-course epidemiology literature^27^ to evaluate the circumstances under which childhood socioeconomic adversity associates with DNAm changes at age 7: 1) *accumulation* hypothesis, where the impact of low SEP increases with the number of time periods exposed, regardless of when it occurs; 2) *sensitive period* hypothesis, where the impact of low SEP is larger in magnitude during a certain developmental period compared to any other; and 3) *mobility* hypothesis, where the impact of SEP on DNAm is driven by an upward or downward change in SEP between adjacent developmental time periods. To our knowledge, this is the first study to simultaneously test these three hypotheses.

Uncovering the dynamic relationships between SEP and DNAm across childhood will not only highlight the biological mechanisms driving the effects of SEP on long-term health, but also will offer clearer insights to guide targeted interventions. From a public health perspective, understanding when (e.g., sensitive periods) and under which circumstances (e.g., downward mobility, or accumulation) the socioeconomic environment is most likely to become biologically embedded can help inform policies that offer safety nets to vulnerable families when the effects of socioeconomic-related adversity might be particularly pernicious for child development.

## Methods

### Sample and procedures

Data came from the Accessible Resources for Integrated Epigenomics Studies (ARIES)^28^, a subsample of 1,018 mother-child pairs from the Avon Longitudinal Study of Parents and Children (ALSPAC). ALSPAC is a prospective, longitudinal birth-cohort in the United Kingdom (UK) designed to investigate genetic and environmental determinants of health across the lifespan^29–31^. Women living in the county of Avon, UK with estimated delivery dates between April 1991 and December 1992 were invited to participate. We analyzed data from 946 singletons in ARIES with blood-based DNAm profiles generated at age 7. Please note the ALSPAC website contains details of all the data available, through a fully searchable data dictionary and variable search tool (http://www.bristol.ac.uk/alspac/researchers/our-data/). Ethical approval for the study was obtained from the ALSPAC Ethics and Law Committee and the Local Research Ethics Committee.

### Measures

#### Early-life socioeconomic position (SEP)

We analyzed six SEP indicators, spanning financial, occupational, and residential domains: (1) job loss, (2) income reduction, (3) low family income, (4) financial hardship, (5) major financial problems, and (6) neighborhood disadvantage. These SEP indicators were measured repeatedly via maternal report through mailed questionnaires during three developmental time periods (**Figure 1a**): *very early childhood* (0-2 years), *early childhood* (3-5 years), and *middle childhood* (6-7 years). These time periods are consistent with previous research studies^21, 32–34^ and roughly correspond to three major developmental stages (infancy/toddlerhood, pre-school, and school-age) when different types of early-life policies and interventions could occur.

**Figure 1.**
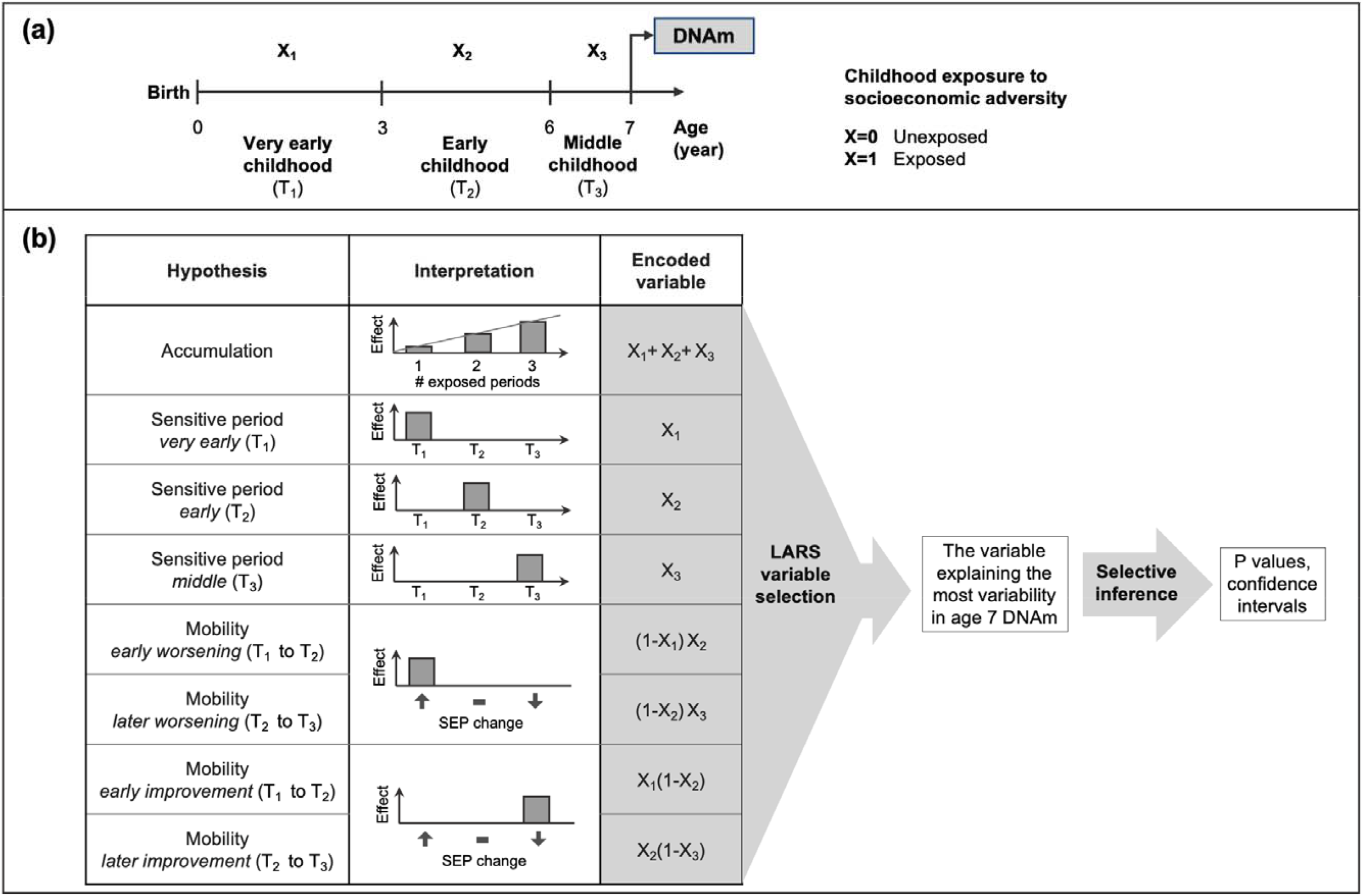
Study design and the conceptual life-course models used in the structured life course modeling approach (SLCMA). (a) Measurement of childhood socioeconomic adversity (X) and DNA methylation (DNAm) over time (T). Exposure to socioeconomic adversities, or indicators of low socioeconomic position (SEP), were measured repeatedly across three childhood periods: very early (0-2 years, T_1_), early (3-5 years, T_2_), and middle childhood (6-7 years, T_3_). DNAm was measured around age 7. (b) Illustration of the life-course hypotheses tested in the SLCMA, the least angle regression (LARS) variable selection procedure, and selective inference test. *Accumulation, sensitive period*, and *mobility* hypotheses were examined in this study. *Accumulation* assumes that the effect of low SEP increases with the number of exposed periods. *Sensitive period* assumes that low SEP is particularly impactful during one of the three time periods. *Mobility* assumes that changes in SEP across specific periods is particularly impactful. *Early worsening* and *early improvement* refer to adversity getting worse (↓SEP) or better (↑SEP) from very early to early childhood, respectively; *later worsening* and *later improvement* refer to adversity getting worse or better from early to middle childhood, respectively. For each socioeconomic adversity, hypotheses were encoded into variables and then entered into the LARS variable selection procedure to identify the one explaining the most variability in DNAm at age 7 at each CpG site. We then performed post-selection inference to test the association between the selected variable and DNAm as well as estimate confidence intervals. See **Supplemental Methods** for more details about SLCMA.

For each SEP indicator, children were classified as exposed or unexposed at each period, using criteria described in **Supplemental Methods**. With these repeated, self-reported SEP indicators, we could identify changes occurring *between* time-periods for indicators capturing time-varying status of SEP. For job loss and income reduction, the measures inherently captured change *within* a certain developmental period, because they asked about socioeconomic mobility. To distinguish job loss and income reduction from other indicators, we refer to them throughout the manuscript as “instability indicators”.

#### DNA methylation (DNAm)

DNAm was measured from peripheral blood at age 7 using the Illumina Infinium HumanMethylation450 BeadChip microarray (Illumina, San Diego, CA). Consent for biological samples was collected in accordance with the Human Tissue Act (2004). DNAm wet laboratory procedures, preprocessing analyses, and quality control have been described elsewhere^28, 35^. A total of 412,956 CpGs on autosomal chromosomes passed quality control and were included in this analysis. For each CpG, DNAm level is expressed as a ‘beta’ value (β-value) ranging from 0 to 1, which represents the proportion of cells methylated at each interrogated CpG. Proportions of the six white cells in the whole blood (CD8 T cells, CD4 T cells, NK cells, B cells, monocytes, and granulocytes) were estimated using Houseman’s method^36^. Estimated cell proportions were included in all analyses to correct for cell type heterogeneity.

#### Covariates

To adjust for baseline demographic differences in ARIES, we controlled for the following variables measured at birth (**Supplemental Methods**) in all analyses: child race/ethnicity, child sex, child birthweight, maternal age, number of previous pregnancies, and sustained maternal smoking during pregnancy. Because age is a strong predictor of DNAm^37^ and the actual time of blood draw at the age 7 assessment varied across children, we also adjusted for child age in months at the time of blood draw (ranging from 85 to 109 months, median=89 months).

### Data analysis

#### Structured life course modeling approach

We used the two-stage structured life course modeling approach (SLCMA)^38–40^ to evaluate the time-dependent effects of socioeconomic adversity on DNAm. SLCMA is a method that leverages repeated exposure data to simultaneously investigate the relationship between exposure and outcome under multiple a priori-defined life-course hypotheses. In our analysis, we tested three life-course hypotheses, described previously, which were parameterized as follows (**Figure 1b**).

First, to test the *accumulation* hypothesis, we created a sum score (ranging from 0 to 3), which captured the number of time periods across the three developmental stages that children were exposed. Second, to test the *sensitive period* hypothesis, we created three binary variables, one for each of the three developmental periods, to classify children’s exposure status (0=child was unexposed during the period; 1=child was exposed during that period). Third, to test the *mobility* hypothesis, we created a pair of indicator variables for change in SEP between very early and early childhood, and a pair of indicator variables for change in SEP between early and middle childhood. Each pair consisted of an indicator variable for worsening (1=change from unexposed to exposed, 0=other) and an indicator variable for improvement (1=change from exposed to unexposed, 0=other). We tested all three hypotheses for low family income, financial hardship, major financial problem, and neighborhood disadvantage. Only the *accumulation* and *sensitive period* hypotheses were tested for job loss and income reduction, the two instability indicators that inherently reflect change (**Table S1**).

We performed the SLCMA in two stages: (1) life-course hypothesis *model selection* followed by (2) *post-selection inference* (**Figure 1b, Supplemental Methods)**. In the first stage, we entered the variables corresponding to the tested hypotheses described above into a Least Angle Regression (LARS) variable selection procedure^41^ to identify the variable explaining the most variability in DNAm. We focused only on the first variable selected to maximize statistical power and prioritize parsimonious explanations^38^. The variable selected represents the life-course hypothesis most supported in the observed data. In the second stage, we used selective inference^38, 42^ to test the association between the variable selected in the first stage and its link to DNAm, as well as estimate confidence intervals. We controlled for baseline demographic variables, child age at blood draw, and cell type proportions in both stages of the analysis.

#### Defining CpG loci of interest

We used two thresholds to identify associations between SEP and CpG loci for further investigation. Given recent recommendations discouraging the use of p-values alone for statistical inference^43, 44^, we used an effect-size-based threshold of R^2^ >3%, meaning that the SEP exposure explained more than 3% of the variance in DNAm. This cutoff was selected based on the effect sizes observed in previous epigenome-wide analyses of childhood adversity in ALSPAC^21, 23^ and other well-established environmental exposures, including tobacco smoking^45^. To maintain consistency with prior literature, we performed multiple-testing correction using the Benjamini-Hochberg method^46^ – a commonly-used method for high-dimensional hypothesis testing – at a 5% false discovery rate (FDR) to assess the statistical significance of top loci.

#### Sensitivity analyses

We conducted three sensitivity analyses to evaluate the robustness of our SLCMA results (**Supplemental Methods**). First, to evaluate the possibility that there might be remaining distortions in the identified SEP-DNAm associations (or residual bias), we additionally controlled for time-invariant SEP indicators (e.g., maternal education at baseline), population substructure estimated from epigenetic data, cord blood DNAm, and genetic variation (at methylation quantitative trait loci, or mQTL). We did not include these variables in main analysis, because prior studies have shown that covariate adjustment may substantially shift the results of DNAm-based analyses^21^; thus, a stepped approach could enable better detection of signal when the role of the covariate in the SEP-DNAm association is unclear, and avoid reducing sample size based on missing covariate data. Second, as *mobility* had never been previously tested on DNAm within childhood to our knowledge, we assessed the additional insights gained by adding *mobility* hypotheses through re-analyzing the CpGs with an R^2^ >3% for low family income, financial hardship, major financial problem, and neighborhood disadvantage using only *accumulation* and *sensitive period* hypotheses. Third, to evaluate the loss (or gain) of information from the SLCMA compared to more conventional epigenetic approaches, we performed an epigenome-wide association study (EWAS) of *any exposure* to each type of SEP adversity before age 7 (0=never exposed; 1=exposed to that type of SEP adversity) and DNAm, thus ignoring the timing or change of SEP over time. See **Supplemental Methods** for details on the rationale and procedures of the sensitivity analyses.

#### Secondary analyses

To interpret our findings and place them in the context of prior literature, we conducted two secondary analyses. We compared the effect estimates of R^2^ >3% CpGs to those reported in previous SEP-related EWAS studies^19^ (**Supplemental Methods**). We also evaluated the biological significance of our findings by examining the correlation between DNAm in blood and brain tissue for the R^2^ >3% CpGs and testing for the enrichment for genomic features, regulatory elements, and Gene Ontology (GO) terms (**Supplemental Methods**).

## Results

### Socioeconomic adversity was common in the ARIES analytic sample

Children included in our analytic sample were mostly White (97.1%) and from both sexes (49.9% female) (**Table S2**). Job loss was the least reported socioeconomic adversity (11.5% ever-exposed), and income reduction was the most common (73.8% ever-exposed) (**Table 1**). For all six socioeconomic adversities, the prevalence of exposure decreased over time (**Table 1, Figure S1**). The average within-SEP correlation ranged from 0.34 to 0.87 (**Table 1**), suggesting these measures were variable across time. The six SEP indicators were moderately correlated with each other during all three childhood periods (polychoric correlation r_avg_=0.35 at very-early childhood, r_avg_=0.34 at early childhood, r_avg_=0.29 at middle childhood, **Figure S2**), suggesting they captured distinct aspects of the socioeconomic environment.

**Table 1.** Prevalence of exposure to socioeconomic adversity by developmental period in the ARIES analytic sample.

|  | **Job loss**<br>**(N=667)** | **Income**<br>**reduction**<br>**(N=711)** | **Low family**<br>**income**<br>**(N=619)** | **Financial**<br>**hardship**<br>**(N=697)** | **Major financial**<br>**problem**<br>**(N=710)** | **Neighborhood**<br>**disadvantage**<br>**(N=687)** |
| --- | --- | --- | --- | --- | --- | --- |
| **Very early childhood**<br>**(0-2 years)** | 42 (6.3%) | 458 (64.4%) | 95 (15.4%) | 127 (18.2%) | 138 (19.4%) | 83 (12.1%) |
| **Early childhood**<br>**(3-5 years)** | 32 (4.8%) | 220 (30.9%) | 79 (12.8%) | 46 (6.6%) | 69 (9.7%) | 36 (5.2%) |
| **Middle childhood**<br>**(6-7 years)** | 18 (2.7%) | 134 (18.9%) | 55 (8.9%) | 29 (4.2%) | 60 (8.5%) | 29 (4.2%) |
| **Ever-exposed<sup>a</sup>** | 77 (11.5%) | 525 (73.8%) | 130 (21.0%) | 147 (21.1%) | 184 (25.9%) | 98 (14.3%) |
| **Average correlation**<br>**over time<sup>b</sup>** | 0.49 | 0.34 | 0.87 | 0.70 | 0.50 | 0.80 |

The first four rows present the number (%) of children who were exposed to the specific type of socioeconomic adversity at each developmental period or ever exposed throughout the three periods.
<sup>a</sup> Children who were exposed during at least one period were defined as ever-exposed for the specific type of socioeconomic adversity.
<sup>b</sup> Polychoric correlations were presented, characterizing the average correlation over time within the given type of exposure.

### Childhood socioeconomic adversities were associated with differences in DNAm at 62 CpGs

We identified 62 CpGs where exposure to socioeconomic adversity explained more than 3% variance in DNAm (R^2^>3%). Most of the 62 CpGs were linked to the two least commonly-reported adversities in ALSPAC: neighborhood disadvantage (17 loci) and job loss (15 loci, **Table 2**). The remaining 30 CpGs were associated with low family income (13 loci), financial hardship (9 loci), major financial problem (5 loci), and income reduction (3 loci, **Table 2**). On average, exposure to socioeconomic adversity was associated with a 3.2% difference in DNAm levels (absolute effect estimates ranged from 0.1-12.8%), explaining 3.3% of the variance in DNAm across CpG sites (R^2^ ranged from 3.0-4.2%) after controlling for covariates (**Table S3**). Exposure to socioeconomic adversity was associated with lower DNAm at most of these loci (43 out of the 62, **Table S3**). Only four of the 62 CpGs identified using the R^2^ cutoff also passed an FDR<0.05 significance threshold, all of which were associated with neighborhood disadvantage (**Table 2**).

**Table 2.** Summary of the SLCMA results for the 62 CpGs with R^2^>3%.

| Adversity | Number of<br>$R^2 > 3\%$ CpGs | Range of $R^2$ | Range of p-values | Number of<br>FDR < 0.05 CpGs |
| --- | --- | --- | --- | --- |
| Neighborhood disadvantage | 17 | 3.0-4.2% | $1.3 \times 10^{-7} - 7.1 \times 10^{-6}$ | 4<sup>a</sup> |
| Job loss | 15 | 3.1-3.7% | $5.8 \times 10^{-7} - 8.8 \times 10^{-6}$ | - |
| Low family income | 13 | 3.0-3.8% | $1.7 \times 10^{-6} - 2.5 \times 10^{-5}$ | - |
| Financial hardship | 9 | 3.0-3.7% | $5.9 \times 10^{-7} - 8.5 \times 10^{-6}$ | - |
| Major financial problem | 5 | 3.0-3.8% | $2.6 \times 10^{-7} - 4.7 \times 10^{-6}$ | - |
| Income reduction | 3 | 3.0-3.3% | $1.5 \times 10^{-6} - 4.5 \times 10^{-6}$ | - |

SLCMA = structured life course modeling approach; FDR = false discovery rate.
<sup>a</sup> Four CpGs for neighborhood disadvantage passed an FDR < 0.05 significance threshold: cg20102336, cg08638097, cg23405172, and cg14212190.

Of note, 61 of these CpGs showed the same direction of positive or negative effect as that reported in at least two prior EWASs examining SEP and DNAm, with 43 loci showing the same direction in five or more prior EWAS analyses. Furthermore, 17 out of the 62 CpGs showed a p<0.05 in at least two prior EWASs, and two CpGs (cg23685969 and cg19260606) exceeded a significance threshold of FDR<0.05 in at least one prior EWAS (**Table S4, Figure S3**).

### Mobility and sensitive period hypotheses were most often selected

Of all life course hypotheses tested, *mobility* and *sensitive period* effects showed the strongest associations with DNAm (**Figure 2a**).

**Figure 2.**
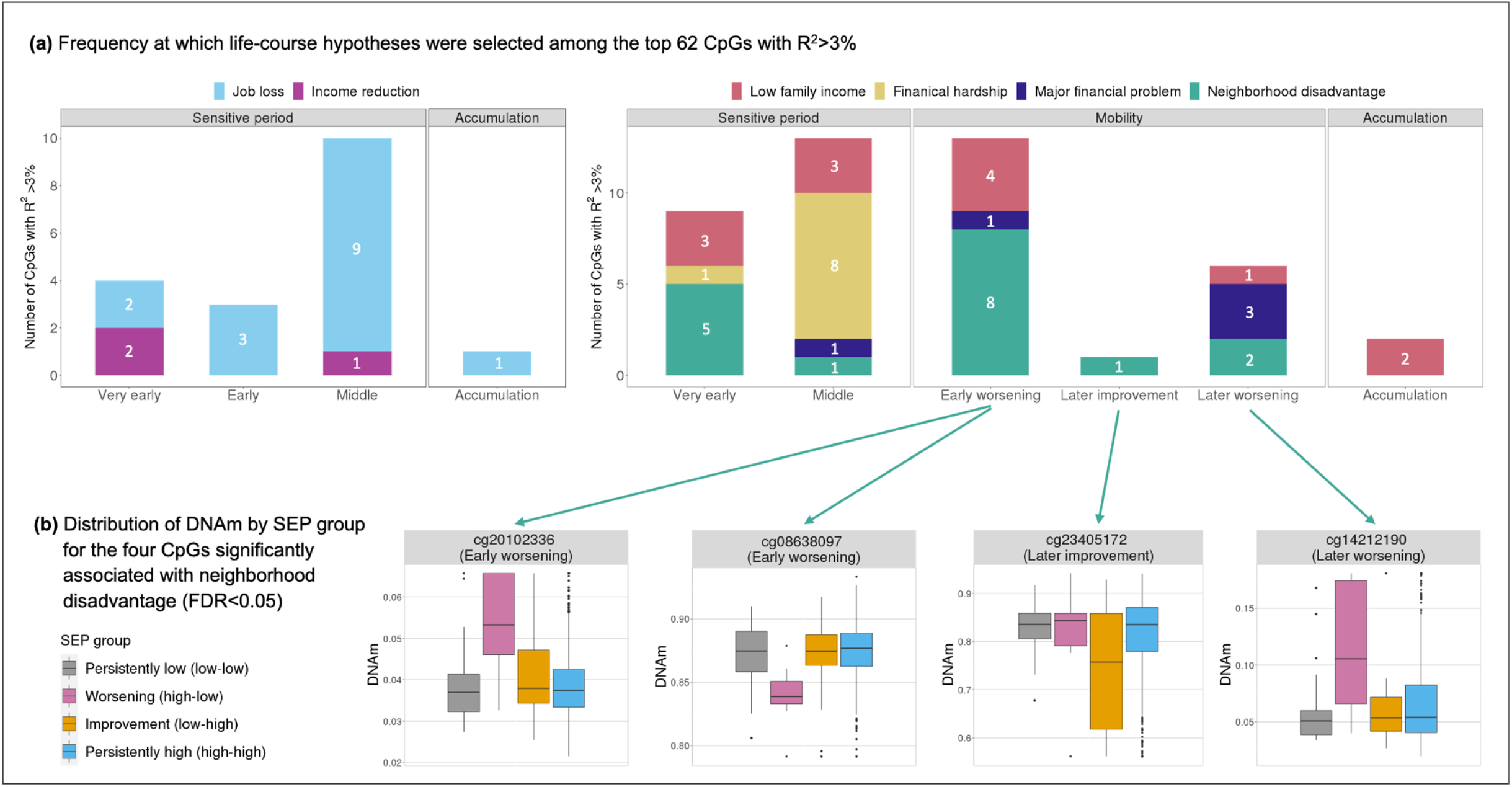
Mobility and sensitive period hypotheses were most often selected among the top 62 CpGs linked with socioeconomic adversity (or socioeconomic position, SEP) that explained > 3% variance in DNA methylation (DNAm). **(a)** Frequency at which each life-course hypothesis was selected among the 62 CpGs. For job loss and income reduction, we tested *accumulatiwon* and *sensitive period* hypotheses, and middle childhood was the most selected hypothesis. For the other four socioeconomic adversities, we tested *accumulation, sensitive period*, and *mobility* hypotheses. *Mobility* hypotheses, specifically worsening SEP, were most selected. *Very early, Early*, and *Middle* refer to sensitive period hypotheses related to the three childhood periods: very early (0-2 years), early (3-5 years), and middle childhood (6-7 years). *Early worsening/improvement* refer to mobility hypotheses for changes between very early and early childhood, and *later worsening/improvement* refer to mobility hypotheses for changes between early and middle childhood. **(b)** For the four CpGs associated with neighborhood disadvantage at an FDR<0.05, SEP mobility group implied by the selected mobility hypothesis showed the greatest shift in DNAm. SEP mobility group was defined based on the exposure status at two consecutive childhood periods (*very early* and *early*, or *early* and *middle*) involved in the mobility hypothesis chosen for each CpG; *persistently low SEP* was defined as being exposed during both periods; *worsening SEP* was defined as being unexposed during the former period but exposed during the later period; *improving SEP* was defined as being exposed during the former period but unexposed during the later period; *persistently high SEP* was defined as being unexposed during both periods.

For the four socioeconomic adversities where all three life-course hypotheses (*accumulation, sensitive period*, and *mobility*) were tested, 44 CpGs (R^2^>3%) were identified, with the majority reflecting *mobility* (20 loci) and *sensitive period* (22 loci) relationships, respectively. The most selected life-course hypothesis varied by socioeconomic adversity. *Sensitive period* was selected for all nine CpGs identified from financial hardship, with middle childhood selected for eight of them (**Figure 2a**). By contrast, *mobility* (worsening SEP) explained more DNAm variability resulting from neighborhood disadvantage (11 of 17 loci) and major financial problem (4 of 5 loci). The time period when *mobility* had the greatest impact differed across SEP indicators, with very early to early childhood most often selected for neighborhood disadvantage, and early to middle childhood most selected for major financial problem (**Figure 2a**). *Accumulation* was only selected for two CpGs, linked to low family income.

Of note, *mobility* hypotheses were selected for all four FDR-significant CpGs, with a worsening hypothesis (meaning *downward mobility*) selected for three of them (**Table S3**). **Figure 2b** shows, for example, at these three CpGs, children exposed to worsening SEP changes had the greatest shift in DNAm as compared to children with other types of SEP trajectories, including those who had persistently low SEP, worsening SEP, improved SEP, or persistently high SEP.

For our instability indicators (job loss and income reduction), which innately capture the effects of socioeconomic mobility, the strongest evidence was again for *sensitive period* effects, with middle childhood (age 3-5) most selected for job loss (9 of 15 loci) and very early childhood (age 0-2) most selected for income reduction (2 of 3 loci) (**Figure 2a**). *Accumulation* was only selected for one CpG linked to job loss.

The same patterns were found at the epigenome-wide level, with most loci showing most variability in response to adversity from *mobility* and *sensitive periods*, rather than the *accumulation* of exposure across development (**Figure S4**).

### SLCMA results were robust to additional covariate adjustment

Our findings were robust to the adjustment of additional covariates, including time-invariant SEP indicators, population substructure, and cord blood DNAm. Specifically, the life-course hypothesis selected by LARS remained the same for all 62 CpGs with R^2^>3% even after these adjustments were made. Almost all CpGs remained significant at the nominal p<0.05 threshold after adjusting for time-invariant SEP indicators (60 loci), population substructure (61 loci), and cord blood DNAm (61 loci, **Table S5**). The associations between socioeconomic adversities and DNAm were also independent of genetic variation previously linked to significant CpGs **(Table S6**).

### Mobility hypotheses improved our ability to identify CpGs related to SEP changes

Considering only accumulation and sensitive period hypotheses, we were unable to fully detect shifts in DNAm patterns related to changes in socioeconomic environment. When mobility hypotheses in SLCMA analyses were omitted, there were minimal changes to the main results showing effects of *sensitive period* on DNAm (n=22 CpGs), as the same hypothesis was selected with similar effect estimates (**Table S7**). However, for CpGs originally linked to *mobility* (n=20), there were substantial attenuations in the estimated SEP-DNAm associations: sensitive period hypotheses were selected instead, which in turn, showed smaller R^2^ (ranging from 0.04-1.6%) and much larger p-values (ranging from 0.001 to 0.84, **Table S7**). These findings suggest that when the underlying association structure is mis-specified, important DNAm signatures may not be identified.

### EWAS of ever-exposed vs. never-exposed failed to identify time-dependent associations

For 59 of the top 62 CpGs (including the 4 FDR-significant CpGs), the effect estimates from the SLCMA were larger in magnitude than those from EWAS (**Figure S5**). In addition, no CpGs with an FDR<0.05 were identified using EWAS of *any exposure*, meaning ever-exposed vs. never-exposed. These findings suggest the SLCMA was better able to identify developmentally sensitive effects of socioeconomic adversity on DNAm profiles, whereas EWAS might fail to detect signals if the true underlying hypothesis was time-dependent^21^.

### Biological significance of SLCMA findings

#### DNAm at significant loci was mildly correlated across blood and brain

We examined the correlation of DNAm at the top 62 CpGs in blood and brain samples, using data from the Blood Brain DNA Methylation Comparison Tool (http://epigenetics.essex.ac.uk/bloodbrain/)^47^. Overall, DNAm was weakly, but positively, correlated between blood and brain regions (**Table S8**) (prefrontal cortex: r_avg_=0.06; entorhinal cortex: r_avg_=0.10; superior temporal gyrus: r_avg_=0.08; cerebellum: r_avg_=0.09). Some CpGs showed particularly strong correlations between blood and brain (e.g., cg24938210, r=0.78 to 0.81 across brain regions).

#### Distinct biological pathways emerged across SEP indicators

The top 62 CpGs showed no significant differences in distributions of genomic features, CpG island locations, or enhancers, as compared to all tested CpGs (Chi-squared tests p>0.05, **Figure S6**).

Gene set enrichment analysis using p-value-ranked epigenome-wide results^48^ showed that SEP-related DNAm patterns were more likely to occur within or near genes involved in neural system regulation, developmental processes, immune functions, metabolic processes, substance localization, and membrane transport (**Figure S7, Figure S8**). However, there was little overlap observed in the significant GO terms across SEP indicators (**Figure S7**), except for one GO term (morphogenesis of a branching epithelium), which emerged in the enrichment analysis for both financial hardship and major financial problem. These findings suggest different socioeconomic adversities may lead to shifts in distinct biological pathways.

## Discussion

The main finding from this study is that *changes* in the socioeconomic environment may coincide with subsequent changes at a biological level as measured through DNAm signatures. Experiencing a change in the socioeconomic environment, particularly worsening neighborhood quality (i.e., mobility) and parental job loss during middle childhood (i.e., sensitive period), explained, on average, a 3.2% difference in DNAm levels, a magnitude of effect commensurate with other potent environmental factors like tobacco smoking. These patterns were detected even after accounting for other dimensions of the socioeconomic environment, ancestry, DNAm levels at birth, and genetic variation. To our knowledge, this study is the first to evaluate the role of socioeconomic changes within childhood in relation to epigenome-wide DNAm.

Our study extends prior literature on the effects of childhood SEP, providing important new insights about the potential biological embedding of changes in the socioeconomic environment. Numerous epigenome-wide analyses have identified a link between childhood SEP and DNAm across the life course^18, 19^. However, only three studies to our knowledge have examined the relationship between socioeconomic mobility and DNAm^49–51^. Each of these three studies included just two timepoints of SEP measures, one in childhood and another in adulthood, and only assessed DNAm in adulthood. Thus, there is limited understanding of the specific ages when mobility might impact DNAm profiles. Epigenomic processes are widely considered to be temporally dynamic^52^; in particular, our results show that DNAm differences linked to socioeconomic mobility and instability can appear in children as early as age 7, suggesting that DNAm during development may be responsive to early-life SEP changes. Outside of the epigenetic literature, a recent review reported that changes in the socioeconomic environment are a unique predictor of child health outcomes^53^. Indeed, prior non-epigenetic studies focused on other SEP-related outcomes in childhood have shown that an episode of parental job loss may have a larger impact on child health and behavior than stable employment in low-income jobs^54–56^. The developmental literature largely suggests that children benefit from stable, predictable environments and that disruptions or breaks in daily routines (e.g., unpredictable events or environmental instability) can be highly stressful^57–59^, impacting cognitive development and other mechanisms implicated in future risk of health and behavioral problems^60^. Our results provide initial evidence that changes in the socioeconomic environment might play a substantial role in shaping DNAm profiles in childhood. As such, future research should consider socioeconomic changes or instability in the context of alterations to epigenetic patterns and other biological outcomes.

Our results also point to the importance of middle childhood as a potential *sensitive period* when the socioeconomic environment might be particularly impactful. We found more evidence for the importance of the developmental timing of socioeconomic adversity on DNAm rather than its accumulation, meaning that the impact of SEP may not uniformly accumulate over time in childhood, but instead may be more impactful during sensitive periods. These results parallel previous findings from the ALSPAC cohort^21^ and elsewhere^24^, suggesting that sensitive period effects can be detected in the epigenome. SEP plays an important role during school-age years^57, 61^, corresponding to our middle childhood time period, when children begin school. Socioeconomic adversity during school-age years may disrupt other domains of the environment present at this developmental period (e.g., changes in parent-child interactions, school, or afterschool care centers). In turn, the elevated stress from exposure during this potential sensitive period may be more likely to influence biological programming through epigenetic modifications and give rise to cascading behavioral effects.

Consistent with prior epigenome-wide studies^51, 62^, we found little overlap between the top CpGs across SEP domains, suggesting that our SEP indicators captured unique aspects of the socioeconomic environment. These findings provide further evidence that various aspects of the SEP construct may trigger distinct mechanisms that lead to different alterations in DNAm patterns^19, 63^. For example, moving to a disadvantaged neighborhood, or experiencing a perceived shift in neighborhood quality, could reflect changes in accessing certain resources, such as playgrounds or other outdoor spaces (resulting in more time spent indoors)^57^, or increase exposure to environmental toxins known to influence gene expression, such as water or air pollution^64, 65^. Indeed, we found that the DNAm alterations linked to neighborhood disadvantage were more likely to occur in genes related to peroxisomes, which are a key component of the biological response to various environmental pollutants^66^. Additionally, we found that experiences of financial hardship (e.g., difficulty in affording common household necessities like food, clothing, heat, and rent) and income reduction were linked to biological pathways related to diet quality, such as nutrient transport and metabolic processes. Overall, different clusters of biological pathways emerged from the DNAm pattern linked to different SEP domains, suggesting that socioeconomic adversities can affect child health through multiple mechanisms.

Across our six SEP indicators analyzed, the majority of loci (17 of 62) detected were related to neighborhood disadvantage, with 4 of these 17 being the only loci to pass an FDR<0.05 significance threshold. These findings point to the important role that neighborhood-level indicators, including more ubiquitous social and physical exposures that are experienced daily by larger segments of a population, may play in shaping the epigenome during child development. Surprisingly, only a handful of studies have investigated neighborhood-level factors in epigenetic research^67, 68^, despite the general consensus that neighborhoods are an important social determinant of health^69^. Indeed, children’s health and well-being are not simply a function of the individual or family circumstances existing inside the home but also intrinsically related to the larger social and physical contexts in which children grow up^70, 71^. Future research should adopt a multi-level approach to studying how the socioeconomic environment, from family income to neighborhood quality, relates to DNAm changes and other biological outcomes that may be ultimately implicated in child development.

While the current study uncovered many insights into SEP and DNAm associations, a major unanswered question is whether these DNAm changes are adaptive or maladaptive, in both the short- and long-term. Teicher and others have noted that early neurobehavioral changes that occur in response to experiences of childhood adversity often enhance immediate survival at the cost of long-term functioning^72^. Thus, are specific epigenomic fluctuations in the face of family socioeconomic adversity reflective of increased resilience, risk, or both? Although we found DNAm differences when comparing children who were exposed vs. unexposed to socioeconomic adversity, we do not know if these SEP-induced shifts were related to functional or behavioral impairment. Future studies should investigate how these DNAm alterations influence subsequent health and behavioral outcomes. Insights from those studies will be critical to discern whether SEP-related DNAm changes influence children’s vulnerability to disease and other negative health/behavioral outcomes.

Should these DNAm markers of socioeconomic adversity be replicated and identified as harmful (rather than adaptive) to health, our findings suggest at least two paths forward for prevention and intervention. First, our results suggest that children and families might benefit from policies and social programs aimed at minimizing socioeconomic instability, especially for lower-income families who may lack a safety-net to draw from during times of transition, such as job loss^73^. Current and emerging programs to promote socioeconomic stability, such as the Supplemental Nutrition Assistance Program^74^ and the American Families Plan^75^, may help preclude epigenetic and other biological changes from arising due to socioeconomic hardships. Through intervention studies like Baby’s First Years^76^, a randomized control trial evaluating how monthly, unconditional cash gifts to families influence children’s cognitive development, the impact of such social programs on the biological level and their potential downstream health and behavioral effects will be better understood.

Second, prevention programs aimed at promoting socioeconomic stability during childhood might benefit from adopting a multisystemic approach that considers the social determinants of health^77^ at the household, neighborhood, and societal level^78^. At the household-level, parent-child interventions that promote supportive parenting styles or social cohesion during times of instability and/or heightened parental stress may help to foster resiliency in children^78, 79^. In fact, parenting interventions that focus on maternal responsiveness have revealed measurable biological impacts on children’s genome-wide DNAm profiles^80^ and on other biomarkers^81–83^. At the neighborhood-level, after-school programs, community recreational centers, or other community-based interventions^65^ that provide added support/routine to children’s environments might be particularly beneficial to children whose families are experiencing socioeconomic instability. Indeed, a recent review reported that community-based intervention programs mitigated the negative effects of childhood adversity on children’s biological outcomes, including DNAm of stress-related genes^84^. Thus, even if families are unable to temporarily afford such community resources due to periods of transition (e.g., being in between jobs), the structure and benefits these programs provide might warrant families to keep accessing them.

The current study should be interpreted in light of several limitations. First, like other epigenome-wide studies of this sample size, we identified few specific CpGs passing a stringent correction for multiple testing. However, following the recent movement to move beyond p-value thresholds alone^43, 44^, we explored the patterns and implications of SEP-related DNAm profiles among top CpGs passing an effect-size-based threshold. The top CpGs passing this threshold were robust to various sensitivity analyses, and there was consistent evidence for the patterns of loci observed, with the majority showing effects in the same direction as previously published findings and two loci showing significance in other studies after correcting for multiple testing. Nevertheless, the results from individual CpGs should be interpreted with caution and validated in larger samples. Second, our analysis focused on childhood up to age 7. It is unclear how long these differential DNAm patterns persist and whether they are reversible in response to subsequent SEP changes, such as upward mobility, later in life. Future studies with DNAm assessments further out in development are needed to establish whether accumulated SEP disadvantage may emerge in DNAm profiles at later ages, compared to the more immediate effects of socioeconomic changes. Third, because this was a population-based sample, extreme cases of socioeconomic disadvantage were likely underrepresented in the ALSPAC cohort. Our results suggest that more severe forms of adversity may have more potent effects, as we identified most top DNAm loci (32 out of 62) from the two socioeconomic adversities that showed the lowest prevalence (job loss and neighborhood disadvantage). Future research in populations with more diverse SEP distributions capturing the fullest gradient (i.e., extreme poverty) will help fully disentangle the impact of SEP on DNAm patterns. Fourth, the ALSPAC cohort is mostly White, which limits generalizability of these findings to other individuals and populations of non-European descent. Prior studies (see review^85^) show ancestry-related variation in DNA methylation that may lead to differences in gene regulation across populations. Thus, future replication efforts are needed in more diverse and representative populations. Finally, this study was observational and based on self-report measures of SEP, which could have been influenced by reporter bias, wherein participant responses may have been shaped by factors like social desirability or recall biases, leading to over- or under-estimates of observed associations^86^. Self-reporting bias is common among survey/questionnaire data in observational studies, especially with sensitive or private topics like income. However, previous research has shown that individual-level SEP measures like education and income, compared to more objective measures assessed at the census tract-level, can more accurately capture the impact of SEP on a number of health outcomes, such as blood pressure and height^87^. Future randomized experiments will help determine the causal effect of socioeconomic adversity on DNAm.

Despite these limitations, our study has several strengths. Unlike most epigenetic studies assuming a single explanation for the underlying association between socioeconomic adversity and DNAm, we explored several possible life-course hypotheses and allowed the best hypothesis to vary across CpGs, which maximized our statistical power to detect loci influenced by socioeconomic adversity. To this end, we detected more signal from the SLCMA than in a conventional EWAS analysis comparing ever-versus never-exposed to childhood adversity. Further, by considering mobility hypotheses, we identified 20 new DNAm markers associated with SEP changes during childhood that would otherwise have been undetected. Taken together, these findings suggest that investigations limited to the presence versus absence of exposure do not reveal the whole picture of the relationship between SEP and DNAm.

In summary, this study adds to a growing literature suggesting that early-life socioeconomic adversity can leave biological memories in the form of DNAm differences in childhood. Uniquely, our findings on socioeconomic mobility and instability highlight that changes in the socioeconomic environment during childhood may trigger disruptions that alter epigenetic programs, at least in the short-term. Ultimately, these findings will enable researchers to build towards better intervention and prevention efforts aimed at reducing socioeconomic disparities and promoting health across the life course.

## Data Availability

The data for the present study are available through the Avon Longitudinal Study of Parents and Children (ALSPAC). The ALSPAC website contains details of all the data available, through a fully searchable data dictionary and variable search tool (http://www.bristol.ac.uk/alspac/researchers/our-data/).

## Acknowledgements

We are extremely grateful to all the families who took part in the ALSPAC study, the midwives for their help in recruiting them, and the whole ALSPAC team, which includes interviewers, computer and laboratory technicians, clerical workers, research scientists, volunteers, managers, receptionists, and nurses. The UK Medical Research Council and the Wellcome Trust (Grant ref: 217065/Z/19/Z) and the University of Bristol provide core support for ALSPAC. ARIES was funded by the BBSRC (BBI025751/1 and BB/I025263/1). Supplementary funding to generate DNA methylation data which is included in ARIES has been obtained from the MRC, ESRC, NIH and other sources. ARIES is maintained under the auspices of the MRC Integrative Epidemiology Unit at the University of Bristol (MC_UU_12013/2 and MC_UU_12013/8). A comprehensive list of grants funding is available on the ALSPAC website (http://www.bristol.ac.uk/alspac/external/documents/grant-acknowledgements.pdf). This publication is the work of the authors, each of whom serve as guarantors for the contents of this paper. We sincerely thank Dr. Andrew J. Simpkin and Dr. Matthew J. Suderman from University of Bristol, and Dr. Esther Walton from the University of Bath, for valuable early feedback on this project. The authors also thank Brigette A. Davis for her assistance with data preparation for this manuscript.

