## Supplemental Methods and Figures for "Socioeconomic changes predict genome-wide DNA methylation in childhood"

**Sample and procedures**

This current study used data from the Accessible Resources for Integrated Epigenomics Studies (ARIES)^1^, a subsample of 1,018 mother-child pairs from the Avon Longitudinal Study of Parents and Children (ALSPAC). ALSPAC is a prospective, longitudinal birth-cohort in the United Kingdom (UK) designed to investigate the genetic and environmental determinants of health across the lifespan^2-4^. Women living in the county of Avon, UK with estimated delivery dates between April 1991 and December 1992 were invited to participate. Approximately 85% percent of eligible pregnant women (N=14,541) agreed to participate and were enrolled, which resulted in 14,062 live births and a sample size of 13,988 children alive at 1 year of age. Response rates to ALSPAC data collection have been good (75% have completed at least one follow-up).

Mother-child pairs in the ARIES were randomly selected from ALSPAC based on availability of DNA samples across five waves of data collection^1^. We used data from 946 singletons in ARIES with blood-based DNA methylation (DNAm) profiles generated at age 7.

Ethical approval for the study was obtained from the ALSPAC Ethics and Law Committee and the Local Research Ethics Committee. Please note that the ALSPAC study website contains details of all the data that is available through a fully searchable data dictionary and variable search tool (http://www.bristol.ac.uk/alspac/researchers/our-data/).

**Measures**

Early-life socioeconomic position (SEP)

Considering the multidimensional nature of socioeconomic position (SEP) and that different indicators of SEP can have distinct relationships with DNAm^5^, we analyzed six SEP indicators, spanning financial, occupational, and residential domains. These SEP indicators were described below, measured via maternal report at least once in each of the following three developmental time periods: *very early childhood* (0-2 years), *early childhood* (3-5 years), and *middle childhood* (6-7 years). These time periods are consistent with previous ALSPAC and other research studies^6-9^, roughly corresponding to three major developmental stages (infancy/toddlerhood, pre-school, and school-age) where different early-life policies and interventions could occur. For SEP indicators assessed more frequently, we collapsed the timepoints so children were classified as exposed to socioeconomic adversity within the time period if they were exposed on at least one occasion within that given time period (**Figure 1a**). Children were classified as exposed using criteria defined below for each construct.

*1. Job loss.* Mothers indicated job loss for themselves and/or their partner and the extent to which it affected them on six occasions after birth (when the child was age 8 weeks, 8 months, 1.75, 2.75 years, 4 years, 5 years, and 6 years). This indicator was assessed on a Likert-type scale ranging from: 1 = ‘yes & affected me a lot’; 2 = ‘yes, moderately affected’; 3 = ‘yes, mildly affected’; 4 = ‘yes, but did not affect me at all’; 5 = ‘no, did not happen’. At each time point, children were coded as exposed if their mothers reported yes to an event which at least mildly affected the family (corresponding to response options 1 to 3 on a 5-point scale, with a lower score reflecting a greater effect).

*2. Income reduction*. Mothers indicated the extent to which a reduction in income in the previous year affected them on seven occasions after birth (when the child was age 8 weeks, 8 months, 1.75 years, 2.75 years, 4 years, 5 years, and 6 years). This indicator was assessed and coded in the same way as job loss, as noted above.

*3. Low family income.* Mothers reported how much their average take home family income was per week (<£100, £100 - £199, £200 - £299, £300 - £399, £400+) at three occasions after birth (when the child was age 2.75 years, 4 years, and 7 years). Children were coded as exposed if their reported family income was in the two lowest income categories (<£100, £100 - £199). This cut-off is consistent with the official average weekly income threshold (i.e., £190 before housing costs) for households living in relative low income as published by the Department for Work and Pensions in its most recent Households Below Average Income (HBAI) report^10^.

*4. Financial hardship*. Mothers indicated the extent to which the family had difficulty affording the following: 1) items for the child; 2) rent or mortgage; 3) heating; 4) clothing; 5) food. Each of the 5 items was coded on a Likert-type scale (1=not difficult; 2=slightly difficult; 3=fairly difficult; 4=very difficult). These items were completed at five occasions after birth (when the child was age 8 months, 1.75 years, 2.75 years, 5 years, and 7 years). At 1.75 years, 2.75 years, and 5 years, a fifth response option was added to some of the items: 5=paid directly by social security. At 7 years, a fifth response option was added to all five items: 5=don’t pay for this. In order to make categories consistent and comparable across occasions, we estimated the difficulty level on the 4-point scale for participants who selected the fifth option (indicating an undefined level of perceived financial hardship) using multiple imputation. We included all SEP indicators and covariates in the imputation, and assigned the rounded mean based on 51 imputation datasets as the imputed difficulty level. We chose an odd number of imputations in order to compare the imputation results between using the mean or the median as the imputed value, and found little difference between the two approaches. Children were coded as exposed if their mothers reported at least fairly difficult for three or more items at each time point (corresponding to response option 3 to 4 on a 4-point scale, with a higher score reflecting more difficulty).

*5. Major financial problem.* Mothers indicated the extent to which a major financial problem affected them on four occasions after birth (when the child was age 8 months, 2.75 years, 5 years, and 6 years). This indicator was assessed and coded in the same way as job loss, as noted above.

*6. Neighborhood disadvantage*. On four occasions after birth (when the child was age 1.75 years, 2.75 years, 5 years, and 7 years), mothers indicated the degree to which the following were problems in their neighborhood: 1) noise from other homes; 2) noise from the street; 3) garbage on the street; 4) dog dirt; 5) vandalism; 6) worry about burglary; 7) mugging; and 8) disturbance from youth. Response options to each item were: 0=not a problem or no opinion, 1=minor problem, 2=serious problem. Items were summed, yielding scores ranging from 0-16. Children with scores of eight or greater, which generally corresponded to the 95th percentile, were classified as exposed to neighborhood disadvantage.

DNA methylation (DNAm)

As described elsewhere^1, 11^, DNA extracted from peripheral blood (whole blood or buffy coat). DNAm was measured at 485,577 CpG dinucleotide sites across the genome using the Illumina Infinium HumanMethylation450 BeadChip microarray, which captures DNAm variation at 99% of RefSeq genes (17 CpG sites per gene, on average). See Relton et al. for details about the laboratory procedures.

The proportion of molecules methylated at each interrogated CpG site on the array was detected using the microarray. The estimated level of DNAm at each CpG sites was expressed as a “beta” value ($\beta$), defined as the ratio of the intensity measured by the methylated probe and the sum of the overall intensity and a recommended offset value $\alpha$ = 100 ($\beta$ = intensity of the Methylated allele (M) / (intensity of the Unmethylated allele (U) + intensity of the Methylated allele (M) + 100)). The $\beta$ value ranges from 0 (no methylated dinucleotides observed) to 1 (all dinucleotides methylated). Background correction and functional normalization were applied to the raw methylation $\beta$-values using the R-package *meffil*, a pipeline developed by Min and colleagues to remove or minimize the effects of variation due to technical artifacts. Cross-hybridizing probes (both autosomal and XY-binding), polymorphic probes, and probes located in sex chromosomes were further excluded from analysis. A total of 412,956 CpGs on autosomal chromosomes passed QC and were included in this analysis. To limit the impact of extreme values, we winsorized the beta values (i.e., values that represent % methylation) at each CpG site, setting the bottom 5% and top 5% of values to the 5th and 95th quantile, respectively.

Additionally, we estimated the proportions of the six white cells in the whole blood (CD8 T cells, CD4 T cells, NK cells, B cells, monocytes, and granulocytes) using Houseman’s method^12^. Estimated cell proportions were included in all analyses to correct for cell type heterogeneity.

The gene symbol of and distance to the nearest gene to each CpG were obtained from the *FDb.InfiniumMethylation.hg19 package* in R^13^. Genomic features including genomic location, relation to CpG islands (CGIs), and enhancers were obtained from Illumina HumanMethylation450 v1.2 Manifest File (<https://support.illumina.com/array/array_kits/infinium_humanmethylation450_beadchip_kit/downloads.html>).

Covariates

To adjust for baseline demographic differences in the cohort, we controlled in the main analyses for the following variables measured at birth: *child race/ethnicity* (0=non-White, 1=White); *child sex* (0=male, 1=female); *child birth weight*; *maternal age* (0=ages 15-19, 1=ages 20-35, 2=age>35); *number of previous pregnancies*; and *sustained maternal smoking during pregnancy* (0=non-smoker, 1=smoker in two or more trimesters, including the third trimester). Because age is a strong predictor of DNAm^14^ and the actual time of blood draw at the age 7 time-point varied across children, we also adjusted for child age in months at the time of blood draw (ranging from 85 to 109 months, median=89 months).

In secondary analyses, we additionally controlled for time-invariant SEP indicators, population substructure, cord blood DNAm, and genetic variation to evaluate the robustness of our findings (see *Additional covariate adjustments* section below).

**Data analysis**

We used a two-stage structured life course modeling approach (SLCMA)^15-17^ to investigate the time-dependent relationship between socioeconomic adversity and DNAm across three life-course hypotheses. SLCMA was performed in two stages: *model selection* followed by *post-selection inference*.

In the first stage, a set of hypotheses were encoded and entered into a model selection procedure (**Figure 1b**). The three life-course hypotheses tested in this analysis were parameterized as follows.

1) *Accumulation* hypothesis, in which the effect of low-SEP increases with the number of periods exposed, regardless of timing. Formally, for the k^th^ period (k=1 for very early childhood, k=2 for early childhood, k=3 for middle childhood), X_k_=1 if exposed to the specific adversity during the k^th^ period, and X_k_=0 if not exposed; then the accumulation hypothesis was coded as X_1_ + X_2_ + X_3_.
2) *Sensitive period* hypotheses, in which the effect of low-SEP depends on the developmental time period of the exposure. We tested three sensitive period hypotheses, one for each childhood period, and the encoded variable was X_k_ for the k^th^ period (k=1,2,3).
3) *Mobility* hypothesis, in which DNAm is associated with an upward or downward change in SEP between adjacent developmental time periods. We tested two improvement hypotheses and two worsening hypotheses: improvement in SEP between k^th^ and (k+1)^th^ period (k=1,2) was coded as X_k_*(1- X_k+1_); worsening in SEP between k^th^ and (k+1)^th^ period (k=1,2) was coded as (1- X_k_)*X_k+1_.

For each SEP indicator, variables encoding the theoretical hypotheses were entered into the Least Angle Regression (LARS) variable selection procedure^18^ to identify the variable explaining the most variability in DNAm at each CpG (R^2^), meaning the hypothesis most supported by the data. For each CpG, six unique hypotheses were selected, corresponding to each SEP indicator.

In the second stage, we performed post-selection inference to test the association between the selected hypothesis and DNAm as well as estimate confidence intervals. We used selective inference^19^, which was recommended for high-throughput applications of the SLCMA^15^.

We implemented the Frisch-Waugh-Lovell (FWL) Theorem^20-22^ to adjust for covariates: first, encoded hypotheses and DNAm were separately regressed on the covariates; then we applied LARS with residuals from the regressions. It has been shown that this approach gives the same effect estimates as a fully adjusted linear model where covariates are included in the regression model directly^23^, while it can improve statistical power and overcome bias. We performed complete-case analysis, meaning that only samples with non-missing exposures and covariates were included in the analysis.

Sensitivity analyses

We conducted three sensitivity analyses to evaluate the robustness of our top results (R^2^>3%) in the primary analysis.

*Additional covariate adjustments*

We reran the SLCMA analysis for the top CpGs adjusting for additional covariates described below to evaluate the possibility of remaining distortions in the identified SEP-DNAm associations (or residual bias).

1) Time-invariant SEP indicators. We examined how our findings were influenced by the adjustment of SEP indicators that were relatively stable throughout childhood. These time-invariant SEP indicators were moderately correlated to the time-varying SEP indicators examined in the main analysis (average correlation with SEP exposures ranged from 0.20 to 0.47 in absolute values). Further controlling for these indicators allowed us to evaluate the robustness of our findings in response to invariant aspects of the socioeconomic environment. These invariant SEP indicators included: *maternal education* (1=less than O-level, 2=O-level, 3=A-level, 4=Degree or above); *maternal marital status* (0=never married, 1=widowed/divorced/separated, 2=married ); *maternal home ownership* (0=owning, 1=rented); *maternal ever homelessness* (0=no, 1=yes); *Townsend Deprivation Index* (an indicator of neighborhood deprivation via Census data, 0=in the first four quintiles, 1=in the highest quintile). We adjusted for each of the time-invariant SEP indicators in a separate model, and also ran a model adjusted for all of them.
2) Population substructure. To assess if our results were biased by population substructure in the sample, we further adjusted for the top four epigenetic principal components (PCs). Although children included in the ARIES sample are primarily White, subtle substructure in the sample may still possibly bias the analysis. Bias may arise if ancestry is associated with both DNAm pattern (e.g. through genetic factors) and exposure to socioeconomic adversities (e.g. through different geographic locations of ancestors)^24^. Therefore, we inferred population structure based on epigenetic PCs estimated using the EPISTRUCTURE algorithm developed by Rahmani et al.^25^, and further adjusted for the top four epigenetic PCs in a sensitivity analysis.
3) Cord blood DNAm. To examine the possibility of remaining bias by prenatal socioeconomic adversities, we further adjusted for cord blood methylation in the model for the top CpGs. Socioeconomic adversities prior to or during pregnancy may affect DNAm *in utero.* Therefore, DNAm level in cord blood can be a proxy of unmeasured prenatal socioeconomic adversity. Sample collection, laboratory procedures, and quality control for cord blood DNAm are described elsewhere^1, 11^. Methylation beta values were normalized, corrected for cell count heterogeneity, and winsorized to remove outliers following the quality control for age 7 DNAm as described above.
4) Genetic variation. For CpGs associated with any methylation quantitative trait loci (mQTLs), based on a database of mQTLs of the ARIES cohort (<http://www.mqtldb.org/>)^26^, we further controlled for genetic variation at mQTLs linked to our top sites. We downloaded the list of mQTLs at age 7, and filtered the data to our top CpG sites. Children were genotyped using the Illumina HumanHap550 quad chip; imputation was performed to the 1000 Genomes (phase 1, version 3, release Dec 2013) reference population using IMPUTE v2.2.2 (25). Variants were filtered by minor allele frequency (MAF>0.01), Hardy-Weinberg equilibrium (HWE>5x10^-7^), and imputation quality (info>0.8); subjects were filtered by missing genotype rate (missingness<3%) and cryptic relatedness (r<0.1). For each top CpG with four or fewer associated SNPs, we included minor allele dosages as additional covariates. For each top CpG site with more than four associated SNPs, we filtered SNPs by call rate (>99%) and ran a principal components analysis among all SNPs associated with each CpG. The top four principal components were used as covariates to represent genetic variation in the sensitivity analysis.

*Analysis without mobility hypothesis*

While *accumulation* and *sensitive periods* have been previously examined for the association between SEP and DNAm^27^, *mobility* within childhood has never been tested on DNAm. When additional hypotheses are added to the SLCMA, there is a cost in terms of statistical power for a fixed sample size. To better understand the value of adding *mobility* hypotheses in the model selection procedure, which could help guide future analyses, we therefore reran the analyses of top CpGs for low family income, financial hardship, major financial problem, and neighborhood disadvantage using only *accumulation* and *sensitive period* hypotheses. We compared the results for the four SEP indicators from analysis with and without *mobility* tested.

*EWAS of exposed vs. unexposed to socioeconomic adversity*

We also performed epigenome-wide association studies (EWAS) of any exposure before age 7 and DNAm to evaluate the loss (or gain) of information from the SLCMA compared to more conventional approaches. For each SEP indicator, children were coded as ever-exposed (versus never-exposed) if they met the exposure criteria at one or more timepoints by age 7. We compared the detected CpGs in EWAS and SLCMA to determine how the two approaches were different in their findings. A mathematical proof is provided elsewhere ^27^ showing that when the true relationship between exposure and outcome depends on the timing or amount of exposure, a standard EWAS of lifetime exposure is underpowered compared to SLCMA.

Secondary analyses

We also conducted two secondary analyses to interpret our findings and place them in the context of prior literature.

*Compare to prior EWAS*

We compared the result of our R^2^>3% CpGs to those identified from seven previous epigenome-wide DNAm studies on socioeconomic position^28-34^, utilizing individual CpG-level summary statistics reported in a recent review paper^5^. In these seven studies, investigators analyzed indicators spanning five SEP domains: household assets, education, occupation, income, and aggregated composite measures. As some studies analyzed more than one SEP domain or SEP at multiple life stages, we included a total of 17 EWASs from the seven studies. Focusing on the 62 CpGs with R^2^>3%, we then compared our results to these prior EWASs to evaluate: (1) the extent to which our effect estimates were in the same direction; (2) whether they presented statistical evidence in those previous studies at the nominal significance level (p<0.05) and at FDR<0.05 (multiple testing correction was performed within each EWAS separately by the aforementioned review paper).

*Exploring the biological significance of the top loci*

DNAm correlation across blood and brain*.* To examine the relevance of SEP-related DNAm pattern identified in peripheral blood tissues to brain health, we used a publicly available database that compare DNAm in peripheral blood tissue and brain tissue. The Blood Brain DNA Methylation Comparison Tool (<http://epigenetics.essex.ac.uk/bloodbrain/>)^35^ includes DNAm levels in whole blood and four brain regions (prefrontal cortex (PFC), entorhinal cortex (EC), superior temporal gyrus (STG), and cerebellum (CER)) in N = 71–75 matched samples from individuals archived in the MRC London Neurodegenerative Disease Brain Bank. This sample includes both neuropathologically unaffected controls and individuals with variable levels of neuropathology. Pearson correlation r values measuring the blood-brain correlations were retrieved from this database.

Enrichment of genomic features and regulatory elements. We examined whether the locations of the R^2^>3% CpGs were enriched in certain genomic regions (e.g., gene body, UTR regions, etc.), CGI and CGI flanking regions (CGI shore, 0–2 kb from CGI; CGI shelf, 2-4 kb from CGI), and enhancers. Annotations of these features were obtained for all R^2^>3% CpGs from the *FDb.InfiniumMethylation.hg19* package in R^13^. We tested if the genomic features were more common among the R^2^>3% CpG sites as compared to all CpGs tested across the epigenome using Chi-squared tests.

Enrichment of biological pathways. Gene set enrichment analyses were conducted using the methylGSA package in R^36^ to identify important biological pathways indicated by our SLCMA results. Epigenome-wide SLCMA results ranked by p-values were used in the gene set enrichment analyses, and Robust Rank Aggregation^37^ was used to adjust for the number of CpG sites in each gene. Gene Ontology (GO) terms were tested, and Bonferroni method was used for multiple testing correction.

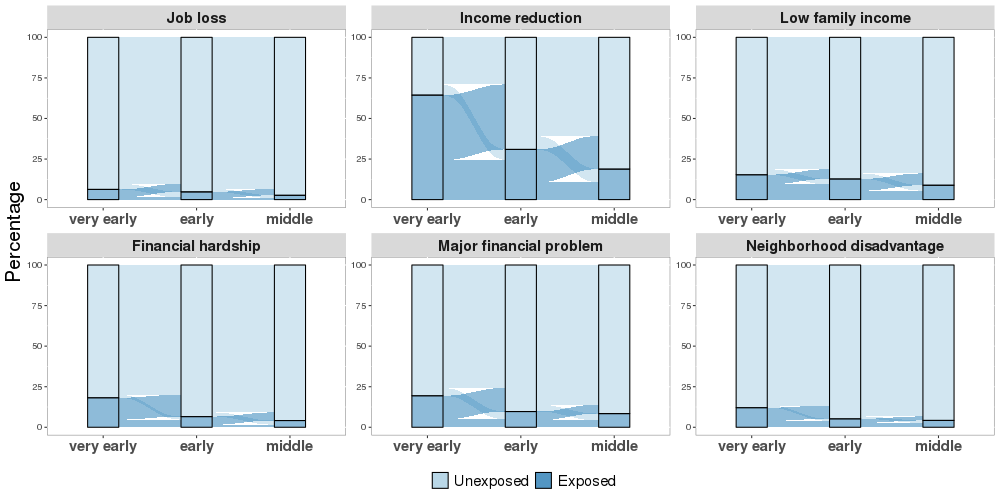

**Figure S1**. Prevalence of each indicator of socioeconomic position (SEP) or socioeconomic adversity, across the three developmental periods. Each panel shows for one of the six SEP indicators the percentage of exposed children (dark blue) and unexposed children (light blue) at three developmental periods as well as the change between periods. For all six SEP indicators, the prevalence of exposure decreased over time; the majority of unexposed children remained unexposed across periods, while exposed children tended to improve in their SEP over time.

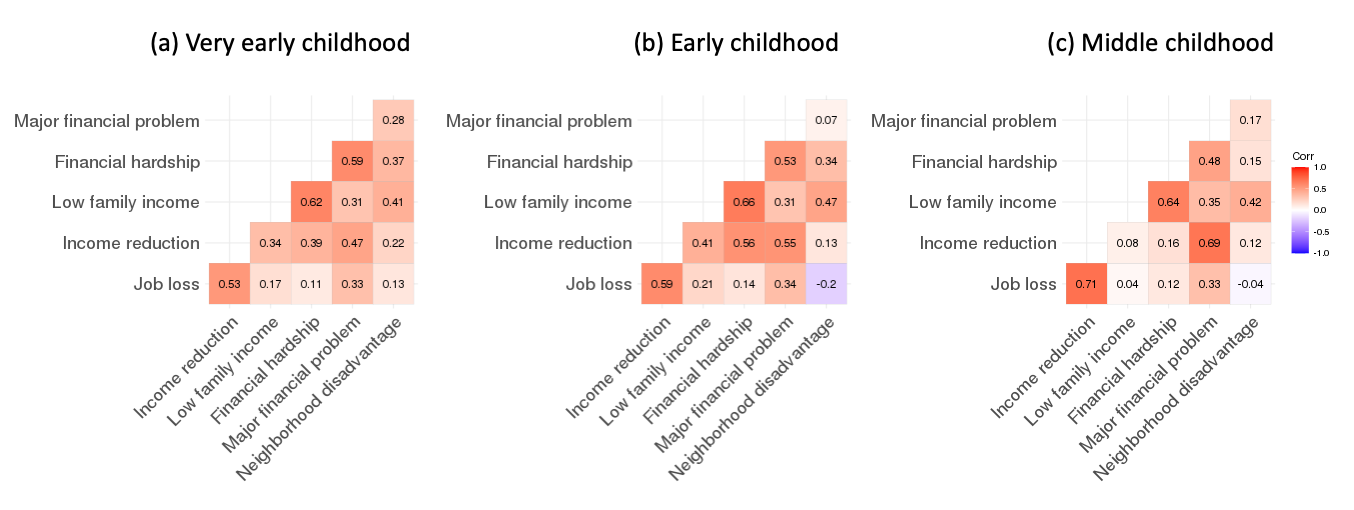

**Figure S2**. Polychoric correlation between socioeconomic adversity pairs during (a) very-early childhood (0-2 years), (b) early childhood (3-5 years), and (c) middle childhood (6-7 years). The six socioeconomic adversities were moderately correlated during all three childhood periods (r_avg_=0.35 at very-early childhood, r_avg_=0.34 at early childhood, r_avg_=0.29 at middle childhood).

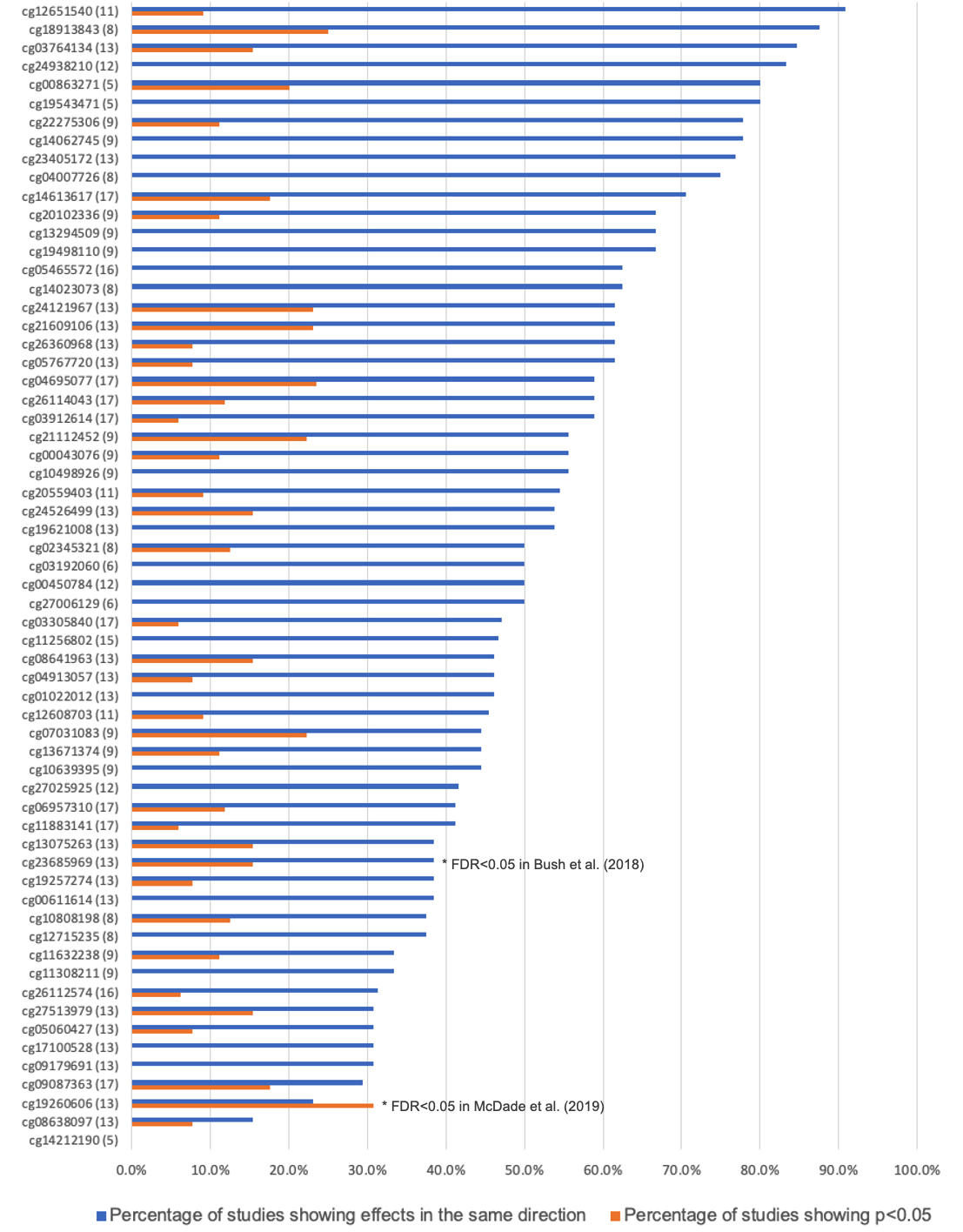

**Figure S3**. Comparison of our results to previous EWAS for the 62 CpGs with R^2^>3%. The x-axis shows the percentage of previous studies showing effects in the same direct (blue) and the proportion of previous studies showing p<0.05 (orange) for each CpG site. The y-axis shows the CpG names with the number of previous EWAS analyses being compared to in the parentheses. Two CpGs passed multiple testing correction at an FDR<0.05 in previous EWAS: cg23685969 was significantly associated with income in Bush et al. (2018) (in our analysis it was significantly associated with low family income); cg19260606 was significantly associated with education and an aggregated composite measure in McDade et al. (2019) (in our analysis it was significantly associated with major financial problem). See **Supplementary Methods** for more details about the studies included.

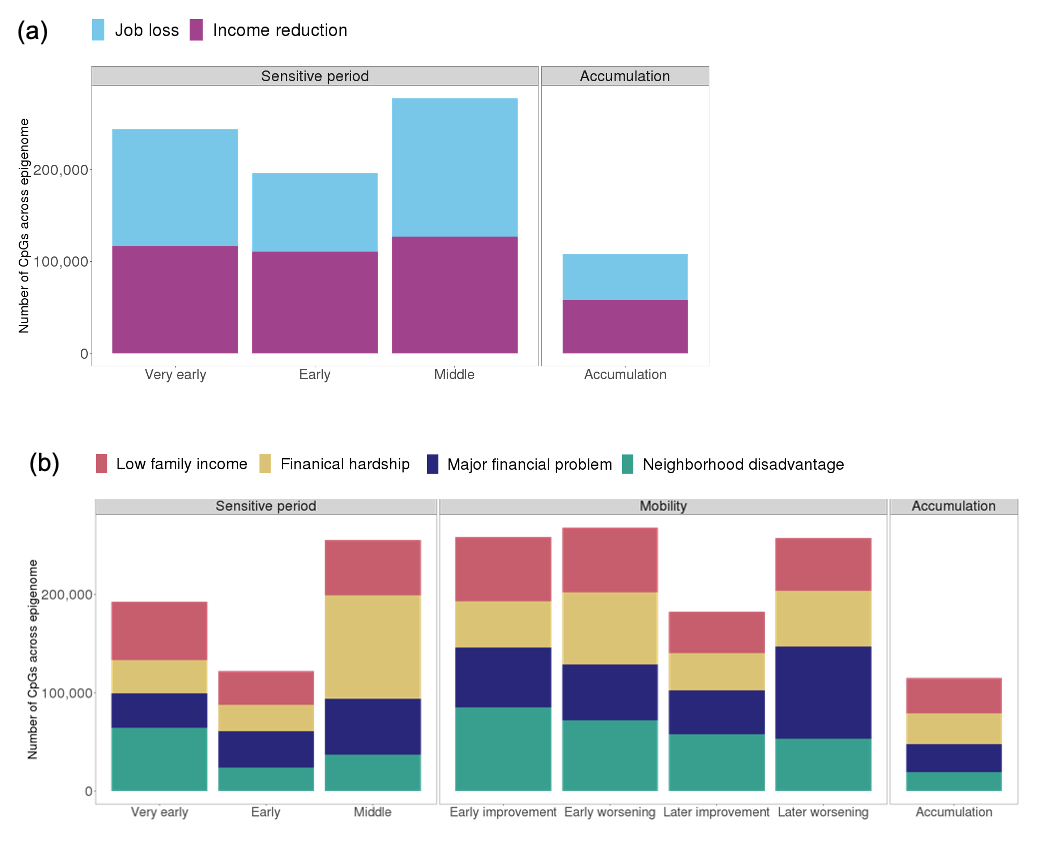

**Figure S4.** Bar plots showing the number of CpGs across the epigenome selected by each life-course hypothesis for each type of socioeconomic adversity. (a) For job loss and income reduction, we tested accumulation and sensitive period hypotheses. (b) For the other four socioeconomic adversities, we tested accumulation, sensitive period, and mobility hypotheses. *Very early*, *Early*, and *Middle* refer to sensitive period hypotheses related to the three childhood periods: very early (0-2 years), early (3-5 years), and middle childhood (6-7 years). *Early worsening/improvement* refer to mobility hypotheses for changes between very early and early childhood, and *later worsening/improvement* refer to mobility hypotheses for changes between early and middle childhood.

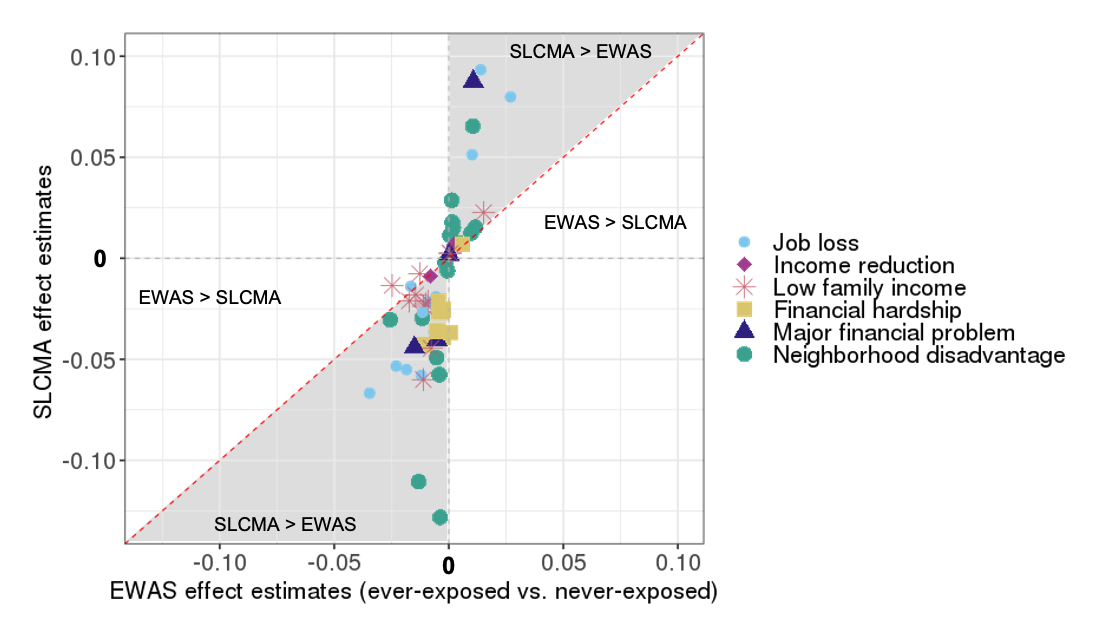

**Figure S5.** Scatterplots showing that the structured life course modeling approach (SLCMA) was more powerful than a standard epigenome-wide association study (EWAS) at identifying time-dependent effect of socioeconomic adversity on DNA methylation (DNAm). This plot compares the effect of neighborhood disadvantage estimated by the SLCMA approach (y-axis) versus those estimated by the standard approach of EWAS (x-axis) of ever-exposure, for the 62 CpGs associated with socioeconomic adversity explaining more than 3% variance in DNAm. The shaded area indicates where the effect estimates were in the same direction in SLCMA and EWAS, but larger in magnitude in SLCMA. The unshaded areas indicates where the effect estimates were greater in the EWAS or estimates were in opposite directions from two analyses. For 59 of the 62 CpGs, the estimated effects were stronger in SLCMA than in EWAS, regardless of the direction of effect.

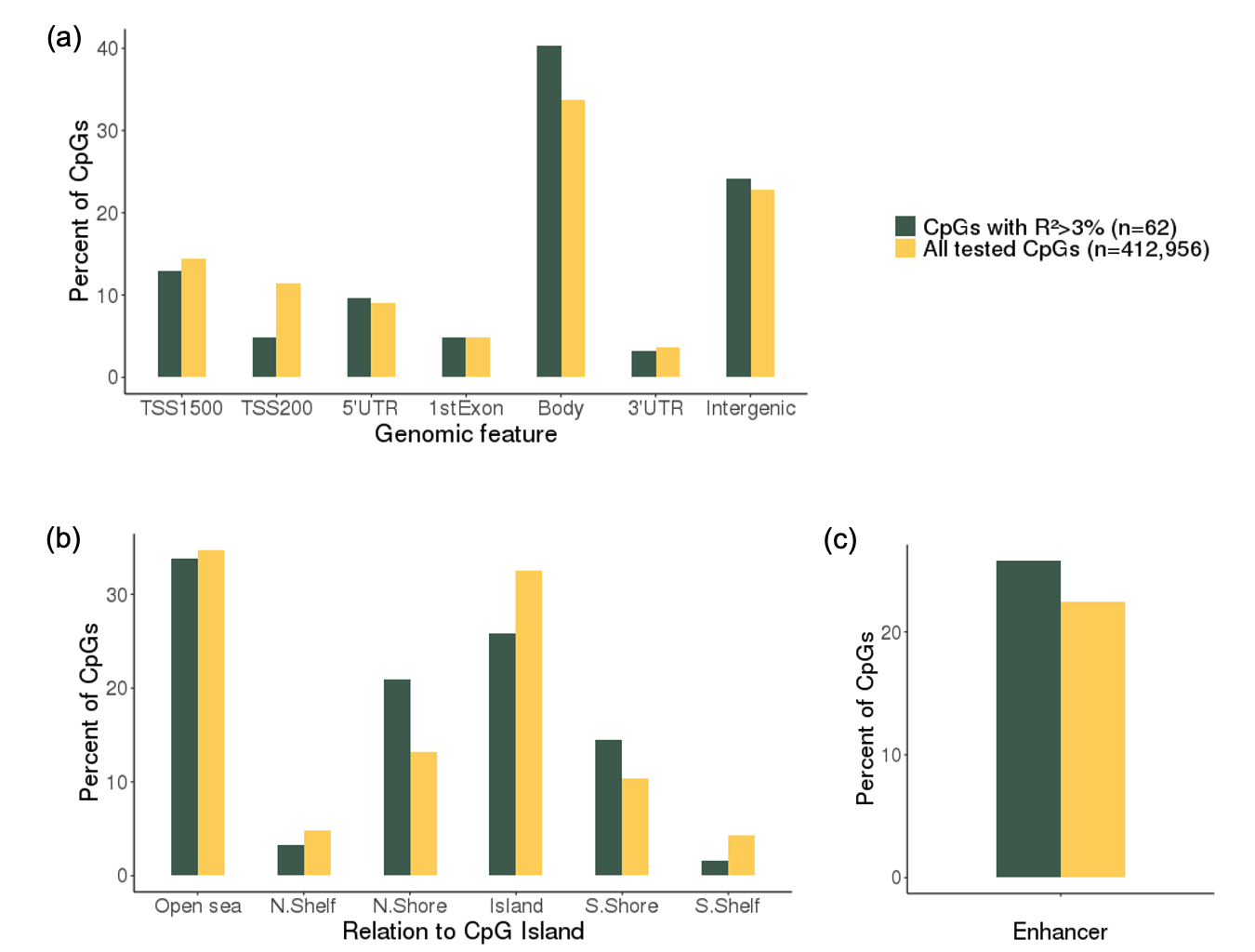

**Figure S6.** The distribution of genomic features (a), CpG island (CGI) locations (b), and enhancer (c) in the 62 CpGs associated with socioeconomic adversity explaining more than 3% variance in DNAm (R^2^>3%, dark green) and all tested CpGs (n=412,956, yellow). TSS1500: within 1500 bp before the transcription start site of a gene. TSS200: within 200 bp before the transcription start site of a gene. CGI shore: 0–2 kb from CGI. CGI shelf: 2-4 kb from CGI.

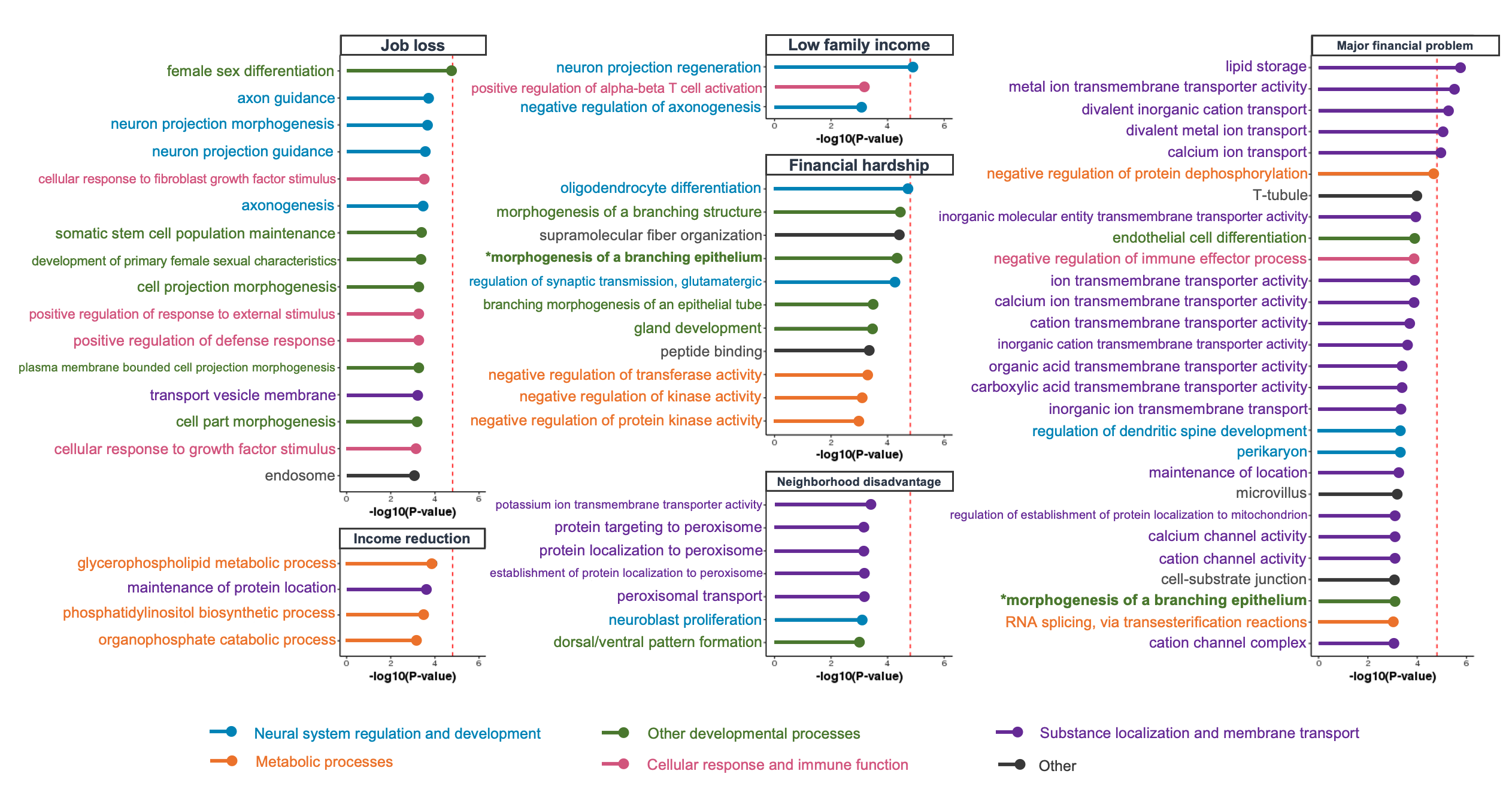

**Figure S7.** Results of gene set enrichment analysis. Gene Ontology (GO) terms that showed p-values<0.001 are shown on the y axis. GO terms were colored by pathway type. The red lines indicate p-value thresholds based on Bonferroni correction. Little overlap in the top pathways was observed across SEP indicators, except for *morphogenesis of a branching epithelium*, which emerged in the enrichment analysis for both financial hardship and major financial problem.

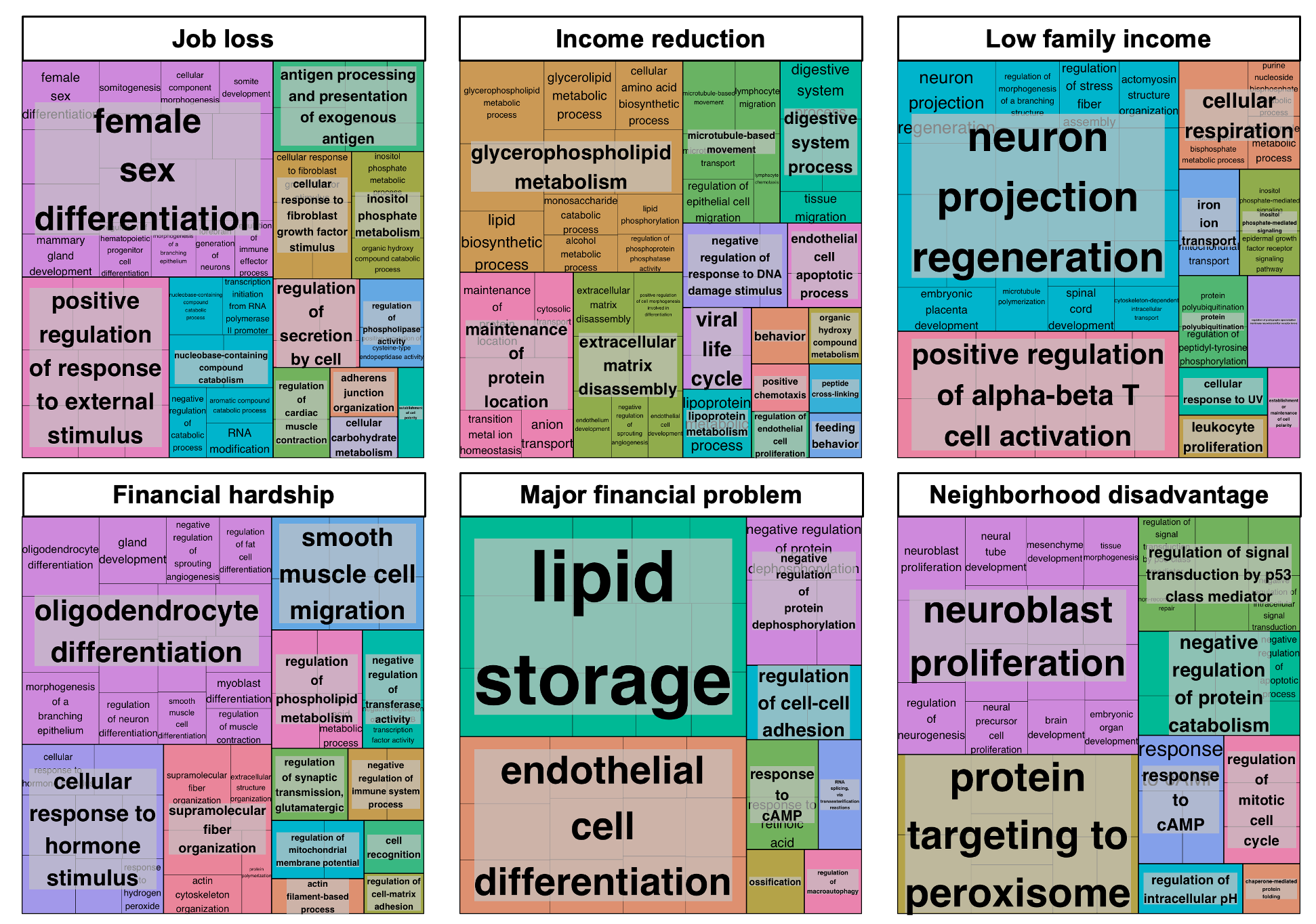

**Figure S8.** Clusters of biological pathways identified in the top 100 Gene Ontology (GO) terms of each SEP measure. GO terms are presented in cells whose size is proportional to the level of significance (-log(p)). Clusters of GO terms were determined by semantic similarity calculated by REVIGO (<http://revigo.irb.hr>). Clusters are labeled by the GO terms with the lowest p value within a cluster.
